# Large-scale osteocyte lacunar morphological analysis of transiliac bone in normal and osteoporotic premenopausal women

**DOI:** 10.1101/2021.12.13.21267731

**Authors:** Elliott Goff, Adi Cohen, Elizabeth Shane, Robert R. Recker, Gisela Kuhn, Ralph Müller

**Affiliations:** Institute for Biomechanics, ETH Zurich, Zurich, Switzerland; Department of Medicine, Columbia University Vagelos College of Physicians & Surgeons, New York, NY, USA; Department of Medicine, Creighton University Medical Center, Omaha, NE, USA

**Keywords:** micro-CT, lacuna, morphometry, osteoporosis, human, osteocyte

## Abstract

Bone’s adaptation ability is governed by the network of embedded osteocytes that inhabit individual crevasses called lacunae. The morphology of these lacunae and their resident osteocytes are known to change with age and diseases such as postmenopausal osteoporosis. However, it is unclear whether alterations in lacunar morphology are present in younger populations with osteoporosis. To investigate this, we implemented a previously validated methodology to image and quantify the three-dimensional morphometries of lacunae on a large scale (26.2 million cells) with ultra-high-resolution micro-computed tomography (microCT) in transiliac bone biopsies from three groups of premenopausal women: control n=39; idiopathic osteoporosis (IOP) n=45; idiopathic low BMD (ILBMD) n=19. Important lacunar morphometric parameters were measured in both trabecular and cortical bone: lacunar density (Lc.N/BV), lacunar porosity (Lc.TV/BV), lacunar number (Lc.N), lacunar volume (Lc.V), lacunar surface area (Lc.S), lacunar alignment (Lc.θ), lacunar stretch (Lc.St), lacunar oblateness (Lc.Ob), lacunar equancy (Lc.Eq), and lacunar sphericity (Lc.Sr). These were then compared against each other and also with previously measured tissue morphometries including: bone volume density (BV/TV), trabecular separation (Tb.Sp), trabecular number (Tb.N), and trabecular thickness (Tb.Th), structure model index (SMI), cortical porosity (Ct.Po) and cortical pore spacing (Ct.Sp). We detected no differences in lacunar morphology between the IOP, ILBMD and healthy premenopausal women. In contrast, we did find significant differences between lacunar morphologies in cortical and trabecular regions within all three groups, which was consistent with our previous findings on a subgroup of the healthy group. Furthermore, we discovered strong correlations between Lc.Sr from both trabecular and cortical regions with the measured BV/TV. The findings and comprehensive lacunar dataset we present here will be a crucial foundation for future investigations of the relationship between osteocyte lacunar morphology and disease.

## Introduction

Osteocytes are the cells within bone tissue that orchestrate bone remodeling via signaling to surface cells [1-5]. They are suspended in fluid and are ensconced within small pores in the mineralized bone matrix, called lacunae. When the bone matrix is compressed by mechanical loading (such as exercise), this results in micro-deformations and fluid-flow shear stress on the osteocyte cell body. The cells then transduce these mechanical signals into chemical ones such as nitric oxide (NO), prostaglandin E2 (PGE2), and bone morphogenic proteins (BMPs) [6, 7]. Consequently, the morphology and distribution of osteocytes and their lacunae is extremely important for the transmitted signal, and the strain concentrations between lacunar structures and extending canaliculi have been shown to drastically amplify the compression experienced at the tissue scale to the cell scale [8, 9]. Therefore, small changes to the lacunar morphology, number of lacunae, and density of the lacunar network could potentially alter the osteocyte’s ability to transduce mechanical signals. Previous studies have demonstrated lacunar morphology differences in patients with low bone mineral density (BMD), such as increased volume and surface area, [10] while others have also shown lower lacunar density in osteoporotic patients relative to control groups [11, 12]. Aging has also been associated with decreased lacunar density, which may contribute to reduced bone remodeling in elderly populations [11, 13-17].

In this study, we investigated the lacunar morphology in iliac crest biopsy samples from premenopausal women with idiopathic osteoporosis (IOP) diagnosed based on history of adult low trauma fracture(s), or idiopathic low bone mineral density (ILBMD) without a history of adult low-trauma fracture, as well as premenopausal healthy non-osteoporotic controls, previously recruited for studies of premenopausal osteoporosis [18-20]. Although most premenopausal women with osteoporosis have a known condition or medication exposure that has caused bone fragility, the affected women recruited for these studies had normal gonadal function and no known disease or medication exposure associated with bone loss. Thus, their rare condition was considered idiopathic. [21].

Previous studies, including biochemistries, histological parameters, and the three-dimensional microarchitecture of transiliac biopsies, in these affected women with idiopathic osteoporosis based on low trauma fracture history (IOP) or very low BMD (ILBMD) have documented thinner, fewer, and more separated trabecular struts relative to the control cohort, as well as thinner cortices [18-20]. Bone remodeling, assessed both by serum bone turnover markers and at the tissue level by transiliac bone biopsy, was quite variable, suggesting that the mechanism of osteoporosis in the affected women may be heterogeneous.

Lacunar morphometric parameters are ideal to investigate as these structures remain intact long after the biopsy has been extracted and the osteocytes have died. Based on these studies, and previous studies reporting differences in lacunae between older osteoporotic subjects and control groups, we hypothesized that differences between control, IOP, and ILBMD groups would also manifest within the 3D morphology of osteocyte lacunae. Additionally, we hypothesized that lacunar morphology would differ between cortical and trabecular regions, as has been previously described [22], as well as correlate with global morphometric parameters such as BV/TV. To test our hypotheses, we imaged and analyzed the same biopsies as the previous studies [18-20], using a validated ultra-high resolution micro-computed tomography (microCT) imaging method to evaluate lacunar morphology in both trabecular and cortical regions [22].

## Methods

### Patient population

Premenopausal women aged 18 – 48 years were recruited at Columbia University Irving Medical Center (New York, NY) and Creighton University (Omaha, NE) with previously reported inclusion and exclusion criteria as described in detail by Cohen et al. [18-20]. To summarize, 104 women were included in this study and composed three groups: healthy control n=40, IOP n=45, and ILBMD n=19. The IOP group included women who had sustained ≥1 low trauma adult fractures irrespective of their areal bone mineral density (aBMD). The ILBMD group included women with low aBMD as determined by dual-energy x-ray absorptiometry (DXA; T score ≤2.5 or Z score ≤ 2.0) who had no history of low trauma adult fractures. Healthy premenopausal women with no adult low trauma fractures and T score /Z score ≥ −1.0 comprised the control group. Secondary causes of osteoporosis were excluded by history and biochemical/hormonal evaluations in all affected subjects and controls. All subjects provided written informed consent and the institutional review boards of both institutions approved these studies.

### MicroCT tissue analysis

Following surgical extraction, fresh biopsies were imaged with a microCT40 (Scanco Medical AG, Brüttisellen, Switzerland) at a nominal resolution of 8μm as previously detailed [18, 23, 24]. An earlier described direct 3D method was implemented to calculate the following trabecular parameters [25]: bone volume density (BV/TV), trabecular separation (Tb.Sp), trabecular number (Tb.N), and trabecular thickness (Tb.Th). Furthermore, the structure model index (SMI) was calculated according to Hildebrand et al. and quantifies the ratio of rod to plate structures within the 3D bone microarchitecture [26]. Cortical indices calculated were cortical porosity (Ct.Po) and cortical pore spacing (Ct.Sp) according to [27].

### MicroCT lacunar analysis

Lacunar imaging and analysis were performed on a microCT50 (Scanco Medical AG, Brüttisellen, Switzerland) at a nominal resolution of 1.2μm as previously described by Goff et al. [22]. Briefly, biopsies were embedded in polymethylmethacrylate (PMMA), cut, and lathed into cylindrical subregions (3.8mm x 10mm) which included both cortical and trabecular regions of each biopsy. Cortical and trabecular 3D image stacks were acquired from each biopsy using optimized scanning parameters: 72 μA current, 4 W power, 55 kVp energy, 1.5 s integration time, level 6 data averaging, and 1500 x-ray projections [22]. Lacunae were segmented via image inversion after applying an individualized threshold based on the distribution of tissue mineral density (TMD) values of each sample. Morphometric parameters of segmented lacunae were calculated with a custom Python script (3.7.1, Python Software Foundation, Delaware, USA) used in combination with XamFlow (Lucid AG, Zürich, Switzerland) and included: Lacunar density (Lc.N/BV), lacunar porosity (Lc.TV/BV), lacunar number (Lc.N), lacunar volume (Lc.V), lacunar surface area (Lc.S), lacunar alignment (Lc.θ), lacunar stretch (Lc.St) and lacunar oblateness (Lc.Ob) as first defined by Mader et al. [28], lacunar equancy (Lc.Eq) as described by Carter et al. [29, 30], and lacunar sphericity (Lc.Sr) as defined by Akhter et al. [31]. Following the nomenclature from Stauber et al. [32], local lacunar parameters normalized to the number of lacunae were denoted as angle brackets (<>) while population lacunar parameters including the entire range of lacunae we labeled with square brackets ([]). The analysis pipeline was previously validated through measures of accuracy, reproducibility, and sensitivity [22]. One biopsy from the control group was excluded because not enough bone was present to scan at high resolution (resulting in n=39).

### Statistical analysis

Student’s t-tests including the required Bonferroni correction factor (p<0.0083) were performed using SPSS (IBM Corp., Version 24.0., Armonk, NY) with respect to the regional inter-group comparison presented in Figure 2. Pearson correlations were performed on the data presented in Figure 3 and significance of correlations were reported in Table 2. Pearson correlations including a linear and quadratic terms were performed on the data in Figure 4.

## Results

### Lacunar morphology

Local and population-based lacunar morphometric indices were measured and compared as presented in Figure 1. Scatter plots contain 103 data points, each representing a local (normalized) lacunar morphometric parameter for an individual patient, while the histograms contain the population-based lacunar morphometric parameters with all 22.6 million lacunae included. Important lacunar morphometric parameters included size dependent measures like volume and surface area (Lc.V and Lc.S) and also slightly more abstract shape tensors like stretch, oblateness, equancy, and sphericity (Lc.St, Lc.Ob, Lc.Eq, and Lc.Sr). Lacunar population distributions appeared to be slightly different between the groups for Lc.TV/BV and Lc.N/BV morphometric parameters, but differences were not significant. Correlations emerged in the scatter plots between parameters such as <Lc.S> and <Lc.V> as well as <Lc.St> and <Lc.Eq>. However, the calculations of these two sets of parameters were inter-related so the correlation was expected. Overall, the lacunar morphometric parameters from the control, IOP, and ILBMD groups were inseparable in the scatter plots in Figure 1. Furthermore, the distribution histograms of the entire lacunar population were indistinguishable among the three groups. This was true for both cortical and trabecular regions. Similar results persisted when IOP and ILBMD groups were combined into a single affected group to compare with controls.

**Figure 1:**
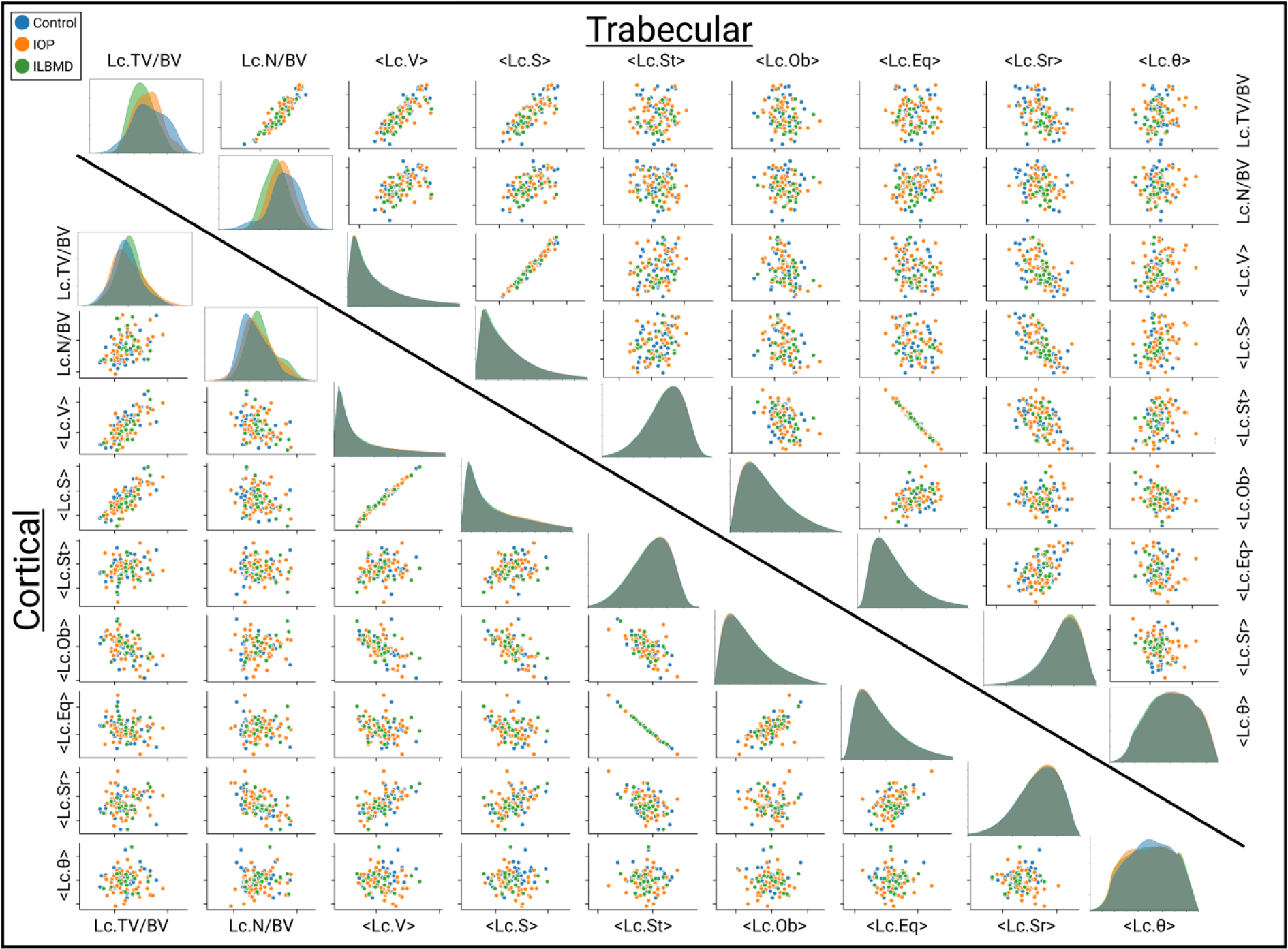
Overview of all measured lacunar morphometric parameters relative to each other. Morphometric parameter distributions and clustering for trabecular regions (top) and cortical regions (bottom). Each plotted point is the mean value of all lacunae from an individual biopsy region (Control, n=39; IOP, n=45; ILBMD, n=19). Histograms display the distribution of the entire lacuna population (n= 22,620,716) from each of the analyzed groups from both regions (Cortical: Control, n=8,371,890; IOP, n=7,206,171; ILBMD, n=3,385,960. Trabecular: Control, n=1,546,019; IOP, n= 1,601,880; ILBMD, n=508,796).

While the three groups were inseparable within the respective trabecular and cortical regions, clear differences emerged between the two regions for each of the three groups for key local lacunar indices such as Lc.N/BV, <Lc.V>, and <Lc.Sr>. Figure 2A-C illustrates significant differences (p<0.001) between cortical and trabecular regions for the control, IOP, and ILBMD groups. Lacunae in cortical regions were significantly larger (<Lc.V>) and more numerous per bone volume (Lc.N/BV) than in trabecular bone. Yet, the lacunae in trabecular bone were significantly more spherical (<Lc.Sr>) than those in the cortical regions. These three differences were consistent for all three groups. The quantitative findings in Figure 2A-C were then qualitatively corroborated by the 3D visualizations presented in Figure 2D-I in which the individual lacunae from the median samples in Figure 2C were colored according to their sphericity. The trabecular lacunae in Figure 2G-I appear overall rounder, sparser, and smaller than in their cortical counterpart displayed in Figure 2D-F. Furthermore, the lacunae visualized in both trabecular and cortical regions appear to be aligned in semi-regular orientations, particularly the less spherical lacunae. Distributions of the ILBMD group appear narrower in Figure 2A-C; however, this may be an artifact present from the recruitment protocol for the group. By definition, ILBMD patients were patients with low bone mass (DXA; T score ≤2.5 or Z score ≤ 2.0) and interestingly a narrower distribution of bone tissue and lacunar morphometric parameters were present in the population as is shown here in Figure 2A-C and further described in Figure 3 Figure 4. Subject characteristics were overall similar between the affected groups as presented in Table 1.

**Figure 2:**
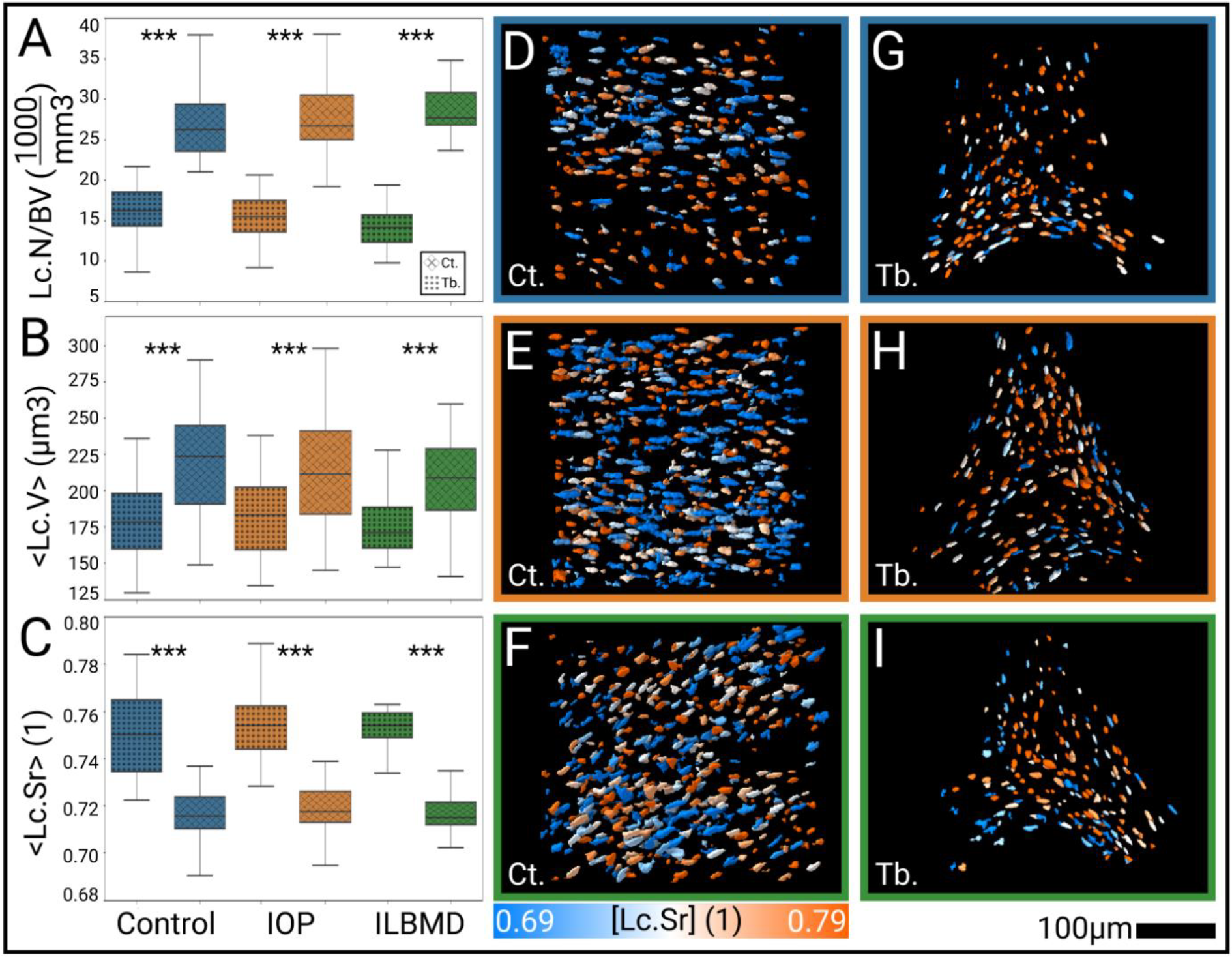
Mean distributions of selected lacunar parameters (A-C) and 3D visualizations of lacunae in both cortical regions (D-F) and trabecular regions (G-I). Local lacunar morphologies compared between region and group for A) lacunar density, B) lacunar volume, and C) lacunar sphericity. Differences between groups were not significant, yet differences between regions within groups were (***=p<0.001). D-F) 3D visualizations of biopsy subsection lacunae from cortical regions colored by [Lc.Sr] for control, IOP, and ILBMD groups respectively. G-I) 3D visualizations of biopsy subsection lacunae from trabecular regions colored by [Lc.Sr] for control, IOP, and ILBMD groups respectively. Biopsies selected for visualization were the median samples from each of the six groups plotted in C). Control (n=39); IOP (n=45); ILBMD (n=19).

**Figure 3:**
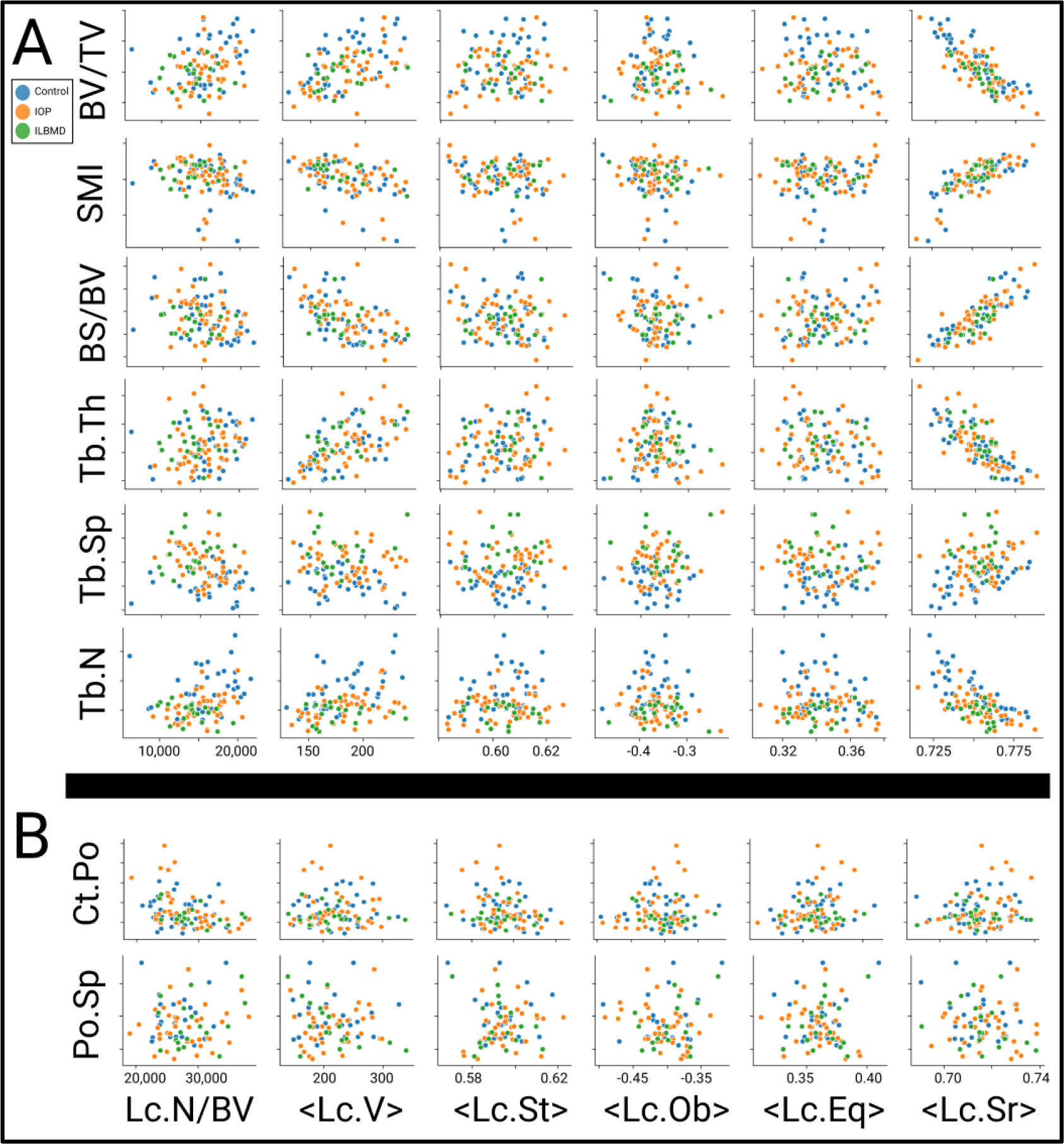
A) Two-parameter comparison between tissue parameters and lacunar morphometric parameters for lacunae from trabecular regions (n=103). B) Two-parameter comparison between tissue parameters and lacunar morphometric parameters for lacunae from cortical regions. Ct.Po measured in the 1.2μm images, while Po.Sp was measured in the 8μm images of biopsies where sufficient cortex was present for evaluation (n=79).

**Figure 4:**
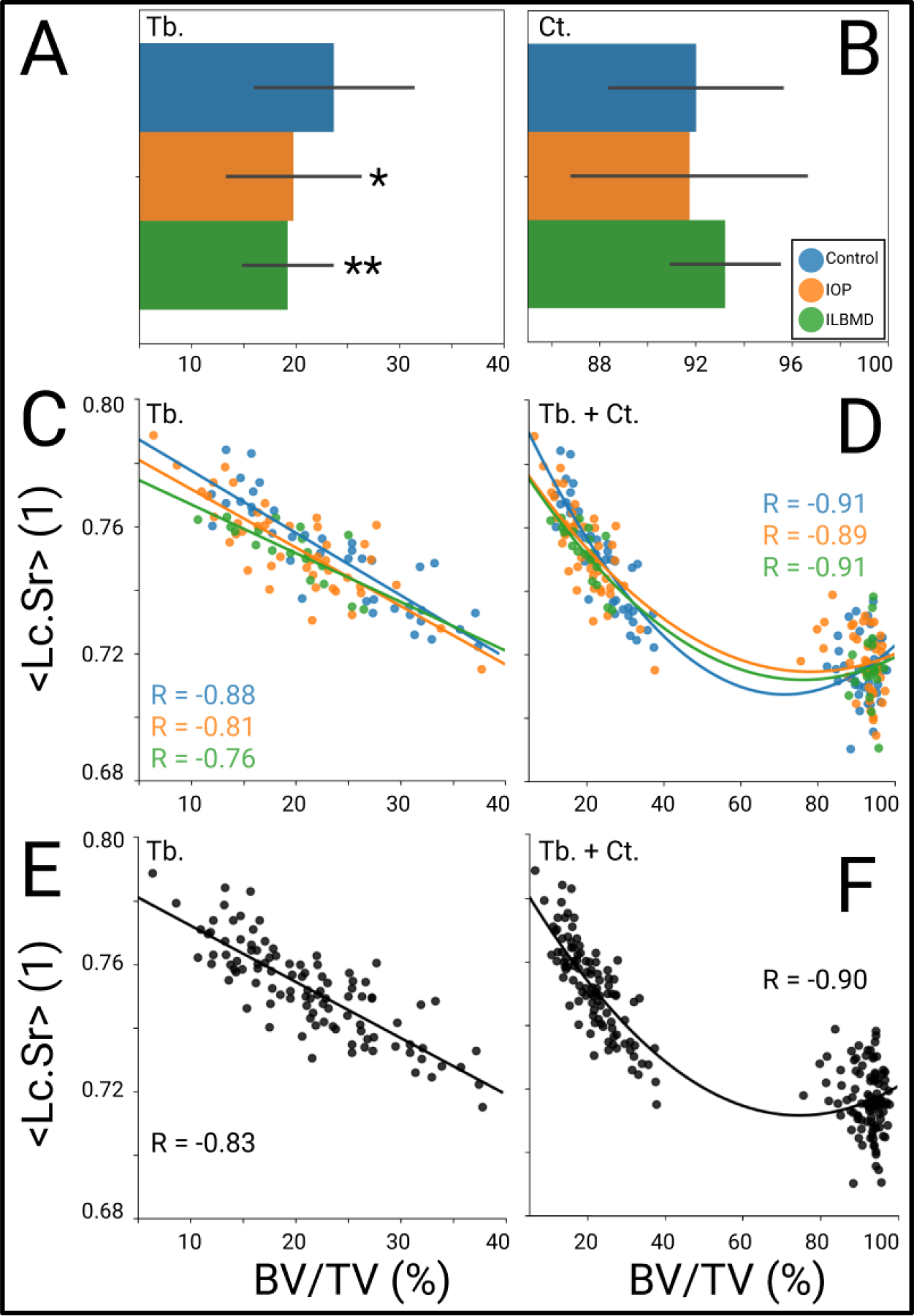
Comparisons and correlations of selected lacunar and tissue morphometric parameters. A) Tissue parameter comparisons between groups for trabecular BV/TV (adapted from Cohen et al. 2011; *=p<0.05, **=p<0.01) and B) cortical BV/TV. C) Linear relationship between <Lc.Sr> and BV/TV in trabecular regions for control (R=-0.88;p<0.001), IOP (R=-0.81;p<0.001), and ILBMD (R=-0.76;p<0.001) groups. D) Quadratic relationship between <Lc.Sr> and BV/TV including lacunae from both trabecular and cortical regions for control (R=-0.91;p<0.001), IOP (R=-0.89;p<0.001), and ILBMD (R=-0.91;p<0.001) groups. E) Linear relationship between <Lc.Sr> and BV/TV in trabecular regions irrespective of group (R=-0.83;p<0.001). F) Quadratic relationship between <Lc.Sr> and BV/TV including lacunae from both trabecular and cortical regions irrespective of group (R=-0.90;p<0.001). Control (n=39); IOP (n=45); ILBMD (n=19).

**Table 1:** Subject characteristics of each group. ^a^ p<0.05 vs. controls, ^b^ p<0.01 vs. controls, ^c^ p<0.001 vs. controls, ^d^ p<0.01 IOP vs. ILBMD.

| Parameter | Control (n=39) | IOP (n=45) | ILBMD (n=19) |
| --- | --- | --- | --- |
| Anthropometric characteristics |  |  |  |
| Age (yr) | 37.5 ± 8.0 | 37.0 ± 7.7 | 39.6 ± 6.3 |
| BMI (kg/m<sup>2</sup>) | 25.5 ± 4.1 | 23.4 ± 4.9<sup>a</sup> | 21.6 ± 3.5<sup>b</sup> |
| BMD (Z score) |  |  |  |
| Lumbar spine | 0.74 ± 0.88 | - 1.33 ± 1.28<sup>c</sup> | - 2.06 ± 0.73<sup>c,d</sup> |
| Total hip | 0.48 ± 0.65 | - 1.09 ± 1.12<sup>c</sup> | - 1.48 ± 0.69<sup>c</sup> |
| Femoral neck | 0.35 ± 0.71 | - 1.31 ± 1.15<sup>c</sup> | - 1.77 ± 0.76<sup>c</sup> |
| Distal radius (one third) | 0.69 ± 0.89 | 0.26 ± 0.82<sup>a</sup> | 0.01 ± 0.85<sup>b</sup> |

### Tissue Morphology

Lacunar morphometric indices for each of the subgroups were plotted against their trabecular and cortical tissue parameters in Figure 3A and B respectively. Each point represents the mean lacunar value from the individual biopsy. Clustering appeared to be tighter visually in the comparisons with trabecular tissue indices when compared to those with the cortical indices but were not significantly different. Apart from the strong and significant <Lc.Sr> correlations in the right-most column in Figure 3A (listed in Table 2), weak correlations were present between several of the trabecular indices and lacunar indices such as volume <Lc.V>; however, none of these were significant.

**Table 2:** Correlations between <Lc.Sr> of lacunae in trabecular regions and measured global trabecular indices. * = p<0.05; **p<0.005; ***p<0.001.

| Tissue Index | Control (n=39) | IOP (n=45) | ILBMD (n=19) | Overall (n=103) |
| --- | --- | --- | --- | --- |
| BV/TV | R=0.88*** | R=0.81*** | R=0.76*** | R=0.83*** |
| SMI | R=0.75*** | R=0.80*** | R=0.71** | R=0.80*** (quadratic) |
| BS/BV | R=0.85*** | R=0.82*** | R=0.72** | R=0.82*** |
| Tb.Th | R=-0.87*** | R=-0.74*** | R=-0.68** | R=-0.78*** |
| Tb.Sp | R=0.54*** | R=0.29 | R=0.01 | R=0.32** |
| Tb.N | R=-0.81*** | R=-0.64*** | R=-0.41 | R=-0.68* (quadratic) |

While no clear separation emerged between the control, IOP, and ILBMD groups, lacunar sphericity evaluated including all three groups was strongly correlated with all respective trabecular parameters as seen in Table 2. All correlations in Table 2 were significant except for the weakest correlations in the Tb.Sp and Tb.N groups. With respect to the cortical regions, Ct.Po was measured on the 1.2μm images while the Po.Sp values reflect the measurements from the 8μm images of which, only 79 biopsies contained a full cortex which was measurable at the tissue level.

BV/TV percentages relative to grouping and the relationship between <Lc.Sr> and BV/TV were examined in greater detail in Figure 4. Figure 4A recapitulates the findings of Cohen et al. that IOP and ILBMD groups had significantly lower BV/TV relative to the control group (p<0.05; p<0.01) [18]. Significant differences did not emerge when comparing the same parameter between groups within the cortical regions in Figure 4B.

Sphericity <Lc.Sr> in trabecular regions was strongly linearly correlated with BV/TV within each group as seen in Figure 4C (R_control = −0.88, p<0.001; R_IOP = −0.81, p<0.001; R_ILBMD = −0.76, p<0.001) and also when evaluated irrespective of grouping in Figure 4E (R = −0.83, p<0.05). Lacunae from cortical regions were included in Figure 4D–F and revealed an even stronger, this time quadratic, correlation between <Lc.Sr> and BV/TV (R_control = −0.91, p<0.001; R_IOP = −0.89, p<0.001; R_ILBMD = −0.91, p<0.001). When evaluated irrespective of grouping, the correlations between <Lc.Sr> and BV/TV was R=-0.90, p<0.001 as presented in Figure 4F.

## Discussion

In this study we investigated osteocyte lacuna distribution and morphology in transiliac biopsies from premenopausal women with IOP and ILBMD and a healthy control group. Contrary to previously published studies of postmenopausal osteoporotic women, lacunar morphology was not altered either in the entire group of affected subjects or when the IOP and ILBMD subgroups were compared to the control group [10-12]. Furthermore, there were no significant group differences with respect to the density of lacunae per bone volume (Lc.N/BV) or the total lacunar porosity (Lc.TV/BV). The inability to distinguish affected and control cohorts based on lacunar morphology was particularly intriguing because these groups differed significantly in previous studies when comparing their 3D microarchitecture, mechanical stiffness, histomorphometry, and biochemistry [18-20]. Yet our data suggest that these differences, which were present at larger spatial scales and in 2D analyses, did not extend to the cellular spatial scale in 3D in these distinct groups of women. These findings also point to the inherent difficulties that are typically associated with idiopathic conditions such as IOP, which may result from heterogeneous mechanisms and etiologies [33], as well as grouping otherwise healthy individuals into a low BMD group (ILBMD) [18]; the only criterion separating the two populations of IOP and ILBMD being an adult low-trauma fracture. This is also a reason to group the IOP and ILBMD subjects into a combined affected group for some analyses.

From a wider perspective, aging has been identified by many studies to be an important driver of osteocyte, lacunar, and canalicular morphology. Busse et al. have shown that lacunar density decreased linearly in humans with respect to age in periosteal and endosteal regions [16], which has been corroborated by additional studies that reflect roughly a 15 to 30% reduction in lacunar density of aged cohorts [11, 14, 15, 34]. Animal studies, notably from Heveran et al., have found similar decreases in lacunar density with respect to age and also discovered lacunar volume to be smaller in old mice [35]. Considering the density of resident osteocytes in humans, a reduction with age has been shown to occur at an even greater pace than the reduction of lacunar density due to a large percentage of empty lacunae [36]. This percentage of empty lacunae has also been shown to increase with age in animal studies (roughly 20 – 30% more in old mice), further highlighting the importance of age in the reduction of viable osteocytes [37-39]. Compounding these two problems in old populations, the dendritic canalicular connections between lacunae have also been shown to decrease with age in both humans and mice [37, 40, 41].

These findings ultimately support the idea that age reduces the embedded network’s functionality from an osteocyte, lacunae, and canalicular perspective. The fact that we find no differences in our lacunar morphometric analyses between affected and control groups of young subjects further strengthens the argument that lacunar changes are age-driven and that we observe our young IOP and ILBMD subjects do not have the characteristic traits of prematurely aged bone at the cellular scale. It is possible that IOP and ILBMD in young women are driven by completely different mechanisms than those causing postmenopausal osteoporosis. The origin of IOP and ILBMD could stem from a wide range of factors such as IGF-I imbalances or reduced osteoblast function, which ultimately may not be associated with morphological changes in osteocyte lacunae at all [42-46].

Within this large dataset of lacunar morphometric indices, strong differences were present between cortical and trabecular bone regions and were apparent both quantitatively and qualitatively as presented in Figure 2. Overall, lacunae were smaller, more spherical, and fewer per bone volume in trabecular bone when compared with cortical bone. Our previous publication detailing the imaging methodology, which was implemented in this study, examined a subset of the control biopsies (n=31) and found the same trends between trabecular and cortical lacunar measured morphometric parameters that we present here [22]. These regional differences extended beyond the healthy cohort and we noted similar differences in both IOP and ILBMD cohorts. The distribution of the lacunar morphometric parameters for the ILBMD cohort (Figure 2A-C) appeared slightly narrower than the other two groups; however, this may be due to the inclusion criteria for the recruitment with narrower DXA BMD entry criteria (DXA; T score <= −2.5 or Z score <= −2.0) [18]. This was further confirmed by the narrow band of clustering of the ILBMD group in the scatter plots of the tissue morphometric indices (Figure 3). Figure 2D-I visually captured the regional differences between trabecular and cortical lacunae that we discovered and displayed the similarity between these lacunar characteristics in healthy, IOP, and ILBMD groups.

We observed strong correlations between <Lc.Sr> and trabecular tissue indices as described in Table 2 and visualized in Figure 3. Although every tissue index we reported is critical for bone microarchitecture quality, many are derived from each other, which also explains the similarities in correlation values in Table 2. BV/TV was the most clinically relevant tissue index we measured since bone mass is broadly used by clinicians to diagnose osteoporosis and osteopenia [47-50]. Therefore, we focused specifically on this relationship in Figure 4, demonstrating the strong, but similar, correlations between groups as well as the overall strong correlation when removing the grouping structure. These correlations between <Lc.Sr> and BV/TV were linear when considering lacunae from trabecular regions (Figure 4C&E). Yet when we included cortical lacunae, which were otherwise uncorrelated with <Lc.Sr> when evaluated alone (Figure 3B), we observed an even stronger, this time quadratic, relationship between <Lc.Sr> and BV/TV (Figure 4D&F). This connection between cortical and trabecular lacunae, which was independent of group (Figure 4F), suggests the possibility that osteocytes remodel their perilacunar space and adjust their lacunar shape similarly depending on the level of BV/TV, regardless of bone’s deterioration at a global level. Furthermore, the quadratic trends we observed in Figure 4D&F emphasizes the importance of this relationship with respect to lacunae in low density, typically trabecular, regions of bone.

Specifically, we observed that lacunae were more spherical in patients with lower BV/TV. This variation in geometry may imply a change in mechanical sensitivity of the lacunar structure, and consequently on the resident osteocyte. Generalizing the range of lacunar shapes to include morphologies between a sphere and a prolate ellipsoid, we can estimate the hoop stress for these ideal geometries. Previously published work in the field of structural geology investigated this topic in porous inorganic material and found that hoop stresses in spherical geometries were roughly twice as high as those in prolate ellipsoids [51]. Applying this finding to our study, we infer that the stresses in spherical lacunae would be generally higher than those in more ellipsoidal lacunae. Furthermore, several previous studies have demonstrated the effect of lacunar stress amplification and its importance for the osteocyte mechanobiology, specifically with respect to the transduction of cell signals [8, 9, 52]. Because of these findings and our correlations in Figure 4, it is a possibility that the osteocytes remodel their direct environment to create more spherical lacunae in regions of weak bone (low BV/TV) as a compensatory mechanism to amplify the mechanical signal. Even though the global remodeling of bone remains impaired, the osteocytes may attempt to correct for this by changing their shape to increase the relative mechanical signal, supporting previous assertions that osteocytes are capable of this type of modulation [10]. This was further supported by the correlation similarities between control, IOP, and ILBMD groups in Figure 4. Perhaps the bone remodeling process would be impaired further without these changes in lacunar sphericity. Yet, in general the changes in the osteocyte lacunar morphology did not appear to be a mechanism for global bone loss in this study, contrary to previously presented histomorphometric evidence in postmenopausal cohorts [13].

The results presented in this study build upon a decade of published findings from the biopsies performed in these premenopausal women [18-20]. The large cohort of healthy individuals (n=39) with which to compare IOP (n=45) and ILBMD (n=19) biopsies demonstrates the power present in our analysis. Within the 103 biopsies, ultimately 22.6 million osteocyte lacunae were measured and morphometrically evaluated. The scale of this lacunar dataset provides a new dimension of information to these biopsies in which the osteocytes were previously only analyzed with histology. Furthermore, our study was performed on a desktop microCT device providing ultra-high resolution and using a newly validated methodology, which illustrates the ability to perform a large-scale osteocyte lacuna imaging study without the need for synchrotron beam line facilities.

Our study also included several limitations, the largest of which being that this was a cross-sectional study and longitudinal data were not available. It is impossible to state definitively if osteocytes remodel their direct environment because of a reduction in BV/TV or if the converse is true: spherical lacunae drive the thinning trend of bone structures at a global level. Both longitudinal animal studies and improved computer simulations will be important to answer this question in future studies [53, 54]. Additionally, specific issues related to the clinical group definitions may have affected the results – since control and ILBMD subjects were recruited based on more narrow BMD criteria. Although the ILBMD group had microarchitectural findings that were quite similar to the IOP group, only the IOP group had a history of low trauma fractures in adulthood leading to greater certainty about a diagnosis of osteoporosis. Furthermore, previous studies have shown that aBMD in small and thin women tends to be lower [55-59]. While the biopsies from our ILBMD group (the group with the lowest BMI) did contain microarchitectural defects [18], it is unknown whether this is an intrinsic characteristic of women with low aBMD and requires a larger population-based longitudinal study to assess further. Younger subjects also may have not yet attained their peak bone mass, and thus lacunar studies may have been affected by developmental changes. Measuring <Lc.Sr> with our previously published imaging method also included some uncertainty with respect to reproducibility (precision error = 0.24%), which manifested in the results we reported [22]. It will also be important for future studies to investigate lacunar morphologies in additional cohorts with different diseases and deficiencies. Perhaps disease driven lacunar changes only occur later in life and are not present in young populations. Additional diseases and hormonal states such as diabetes, osteonecrosis, renal osteodystrophy, and hyperparathyroidism are known to impact skeletal fragility, are associated with bone resorption, and thus, the lacunar morphologies from these populations will be important to compare with our dataset [60-65]. Furthermore, previous studies have suggested that in cohorts of men, IOP pathogenesis is associated with reduced osteoblast function – another important dataset to compare with lacunar morphometric parameters [45, 46].

## Conclusions

Ultimately, we have successfully imaged 26.2 million lacunae in 103 human biopsies, which when coupled with previously measured 3D microarchitecture data, revealed a profound connection between lacunar sphericity and bone volume fraction. We also uncovered significant differences between lacunae in trabecular and cortical regions, which corroborates our previous findings within a subset of the control samples [22]. While we expected to discover differences in lacunar morphologies among the healthy, IOP and ILBMD groups, it is even more interesting to discover how consistent the distributions, trends, and relationships of lacunar morphologies remained across the three groups. Perhaps mechanisms leading to osteoporosis associated with aging are more likely to be associated with changes in lacunar morphology than the mechanisms underlying osteoporosis in younger cohorts with IOP and ILBMD. These findings provide a solid foundation for longitudinal studies, multiscale computer simulations of bone, and comparisons with other disease pathologies in the future.

## Data Availability

All data produced in the present study are available upon reasonable request to the authors

## Acknowledgements

None.

## References

[1] J. Gluhak-Heinrich, L. Ye, L.F. Bonewald, J.Q. Feng, M. MacDougall, S.E. Harris, D. Pavlin, Mechanical loading stimulates dentin matrix protein 1 (DMP1) expression in osteocytes in vivo, J Bone Miner Res 18(5) (2003) 807–17.

[2] A.G. Robling, P.J. Niziolek, L.A. Baldridge, K.W. Condon, M.R. Allen, I. Alam, S.M. Mantila, J. Gluhak-Heinrich, T.M. Bellido, S.E. Harris, C.H. Turner, Mechanical stimulation of bone in vivo reduces osteocyte expression of Sost/sclerostin, J Biol Chem 283(9) (2008) 5866–75.

[3] L. Lanyon, Osteocytes, strain detection, bone modeling and remodeling, Calcified tissue international 53(1) (1993) S102–S107.

[4] F.A. Schulte, D. Ruffoni, F.M. Lambers, D. Christen, D.J. Webster, G. Kuhn, R. Muller, Local mechanical stimuli regulate bone formation and resorption in mice at the tissue level, PLoS One 8(4) (2013) e62172.

[5] R. Huiskes, R. Ruimerman, G.H. van Lenthe, J.D. Janssen, Effects of mechanical forces on maintenance and adaptation of form in trabecular bone, Nature 405(6787) (2000) 704–6.

[6] L.D. You, S. Temiyasathit, P.L. Lee, C.H. Kim, P. Tummala, W. Yao, W. Kingery, A.M. Malone, R.Y. Kwon, C.R. Jacobs, Osteocytes as mechanosensors in the inhibition of bone resorption due to mechanical loading, Bone 42(1) (2008) 172–179.

[7] A. Santos, A.D. Bakker, H.M. Willems, N. Bravenboer, A.L. Bronckers, J. Klein-Nulend, Mechanical loading stimulates BMP7, but not BMP2, production by osteocytes, Calcif Tissue Int 89(4) (2011) 318–26.

[8] A.R. Bonivtch, L.F. Bonewald, D.P. Nicolella, Tissue strain amplification at the osteocyte lacuna: a microstructural finite element analysis, Journal of biomechanics 40(10) (2007) 2199–2206.

[9] A.R. Stern, D.P. Nicolella, Measurement and estimation of osteocyte mechanical strain, Bone 54(2) (2013) 191–5.

[10] R.P. van Hove, P.A. Nolte, A. Vatsa, C.M. Semeins, P.L. Salmon, T.H. Smit, J. Klein-Nulend, Osteocyte morphology in human tibiae of different bone pathologies with different bone mineral density - Is there a role for mechanosensing?, Bone 45(2) (2009) 321–329.

[11] M. Mullender, D. Van der Meer, R. Huiskes, P. Lips, Osteocyte density changes in aging and osteoporosis, Bone 18(2) (1996) 109–113.

[12] S. Qiu, D.S. Rao, S. Palnitkar, A.M. Parfitt, Reduced iliac cancellous osteocyte density in patients with osteoporotic vertebral fracture, Journal of bone and mineral research 18(9) (2003) 1657–1663.

[13] M. Mullender, S.D. Tan, L. Vico, C. Alexandre, J. Klein-Nulend, Differences in osteocyte density and bone histomorphometry between men and women and between healthy and osteoporotic subjects, Calcified tissue international 77(5) (2005) 291–296.

[14] D. Vashishth, O. Verborgt, G. Divine, M.B. Schaffler, D.P. Fyhrie, Decline in osteocyte lacunar density in human cortical bone is associated with accumulation of microcracks with age, Bone 26(4) (2000) 375–380.

[15] S. Qiu, D. Rao, S. Palnitkar, A. Parfitt, Age and distance from the surface but not menopause reduce osteocyte density in human cancellous bone, Bone 31(2) (2002) 313–318.

[16] B. Busse, D. Djonic, P. Milovanovic, M. Hahn, K. Püschel, R.O. Ritchie, M. Djuric, M. Amling, Decrease in the osteocyte lacunar density accompanied by hypermineralized lacunar occlusion reveals failure and delay of remodeling in aged human bone, Aging cell 9(6) (2010) 1065–1075.

[17] B. Busse, M. Hahn, T. Schinke, K. Püschel, G.N. Duda, M. Amling, Reorganization of the femoral cortex due to age-, sex-, and endoprosthetic-related effects emphasized by osteonal dimensions and remodeling, Journal of Biomedical Materials Research Part A: An Official Journal of The Society for Biomaterials, The Japanese Society for Biomaterials, and The Australian Society for Biomaterials and the Korean Society for Biomaterials 92(4) (2010) 1440–1451.

[18] A. Cohen, D.W. Dempster, R.R. Recker, E.M. Stein, J.M. Lappe, H. Zhou, A.J. Wirth, G.H. van Lenthe, T. Kohler, A. Zwahlen, R. Muller, C.J. Rosen, S. Cremers, T.L. Nickolas, D.J. McMahon, H. Rogers, R.B. Staron, J. LeMaster, E. Shane, Abnormal bone microarchitecture and evidence of osteoblast dysfunction in premenopausal women with idiopathic osteoporosis, J Clin Endocrinol Metab 96(10) (2011) 3095–105.

[19] A. Cohen, X.S. Liu, E.M. Stein, D.J. McMahon, H.F. Rogers, J. LeMaster, R.R. Recker, J.M. Lappe, X.E. Guo, E. Shane, Bone Microarchitecture and Stiffness in Premenopausal Women with Idiopathic Osteoporosis, J. Clin. Endocrinol. Metab. 94(11) (2009) 4351–4360.

[20] A. Cohen, R.R. Recker, J. Lappe, D.W. Dempster, S. Cremers, D.J. McMahon, E.M. Stein, J. Fleischer, C.J. Rosen, H. Rogers, R.B. Staron, J. Lemaster, E. Shane, Premenopausal women with idiopathic low-trauma fractures and/or low bone mineral density, Osteoporos Int 23(1) (2012) 171–82.

[21] H.M. Heshmati, S. Khosla, Idiopathic osteoporosis: a heterogeneous entity, Ann. Med. Interne 149(2) (1998) 77–81.

[22] Elliott Goff, Federica Buccino, Chiara Bregoli, Jonathan P. McKinley, Basil Aeppli, Robert R. Recker, Elizabeth Shane, Adi Cohen, Gisela Kuhn, Ralph Müller, Large-scale quantification of human osteocyte lacunar morphological biomarkers as assessed by ultra-high-resolution desktop micro-computed tomography, Bone 152 (2021) 116094.

[23] A. Cohen, D.W. Dempster, R. Muller, X.E. Guo, T.L. Nickolas, X.S. Liu, X.H. Zhang, A.J. Wirth, G.H. van Lenthe, T. Kohler, D.J. McMahon, H. Zhou, M.R. Rubin, J.P. Bilezikian, J.M. Lappe, R.R. Recker, E. Shane, Assessment of trabecular and cortical architecture and mechanical competence of bone by high-resolution peripheral computed tomography: comparison with transiliac bone biopsy, Osteoporosis International 21(2) (2010) 263–273.

[24] P. Ruegsegger, B. Koller, R. Muller, A microtomographic system for the nondestructive evaluation of bone architecture, Calcified Tissue International 58(1) (1996) 24–29.

[25] T. Hildebrand, A. Laib, R. Muller, J. Dequeker, P. Ruegsegger, Direct three-dimensional morphometric analysis of human cancellous bone: Microstructural data from spine, femur, iliac crest, and calcaneus, Journal of Bone and Mineral Research 14(7) (1999) 1167–1174.

[26] T. Hildebrand, P. Ruegsegger, Quantification of Bone Microarchitecture with the Structure Model Index, Comput Methods Biomech Biomed Engin 1(1) (1997) 15–23.

[27] M.L. Bouxsein, S.K. Boyd, B.A. Christiansen, R.E. Guldberg, K.J. Jepsen, R. Muller, Guidelines for Assessment of Bone Microstructure in Rodents Using Micro-Computed Tomography, Journal of Bone and Mineral Research 25(7) (2010) 1468–1486.

[28] K.S. Mader, P. Schneider, R. Muller, M. Stampanoni, A quantitative framework for the 3D characterization of the osteocyte lacunar system, Bone 57(1) (2013) 142–54.

[29] Y. Carter, C.D.L. Thomas, J.G. Clement, A.G. Peele, K. Hannah, D.M.L. Cooper, Variation in osteocyte lacunar morphology and density in the human femur - a synchrotron radiation micro-CT study, Bone 52(1) (2013) 126–132.

[30] A. Carriero, M. Doube, M. Vogt, B. Busse, J. Zustin, A. Levchuk, P. Schneider, R. Muller, S.J. Shefelbine, Altered lacunar and vascular porosity in osteogenesis imperfecta mouse bone as revealed by synchrotron tomography contributes to bone fragility, Bone 61 (2014) 116–124.

[31] M.P. Akhter, D. Kimmel, J. Lappe, R. Recker, Effect of Macroanatomic Bone Type and Estrogen Loss on Osteocyte Lacunar Properties in Healthy Adult Women, Calcified Tissue International (2017) 1–12.

[32] M. Stauber, R. Muller, Volumetric spatial decomposition of trabecular bone into rods and plates - A new method for local bone morphometry, Bone 38(4) (2006) 475–484.

[33] S. Khosla, E.G. Lufkin, S.F. Hodgson, L.A. Fitzpatrick, L.J. Melton, Epidemiology and clinical features of osteoporosis in young individuals, Bone 15(5) (1994) 551–555.

[34] S. Mori, R. Harruff, W. Ambrosius, D. Burr, Trabecular bone volume and microdamage accumulation in the femoral heads of women with and without femoral neck fractures, Bone 21(6) (1997) 521–526.

[35] C.M. Heveran, A. Rauff, K.B. King, R.D. Carpenter, V.L. Ferguson, A new open-source tool for measuring 3D osteocyte lacunar geometries from confocal laser scanning microscopy reveals age-related changes to lacunar size and shape in cortical mouse bone, Bone 110 (2018) 115–127.

[36] S. Qiu, D. Rao, S. Palnitkar, A. Parfitt, Relationships between osteocyte density and bone formation rate in human cancellous bone, Bone 31(6) (2002) 709–711.

[37] L.M. Tiede-Lewis, Y.X. Xie, M.A. Hulbert, R. Campos, M.R. Dallas, V. Dusevich, L.F. Bonewald, S.L. Dallas, Degeneration of the osteocyte network in the C57BL/6 mouse model of aging, Aging-US 9(10) (2017) 2187-+.

[38] M. Piemontese, M. Almeida, A.G. Robling, H.-N. Kim, J. Xiong, J.D. Thostenson, R.S. Weinstein, S.C. Manolagas, C.A. O’Brien, R.L. Jilka, Old age causes de novo intracortical bone remodeling and porosity in mice, JCI insight 2(17) (2017).

[39] L.B. Meakin, G.L. Galea, T. Sugiyama, L.E. Lanyon, J.S. Price, Age-related impairment of bones’ adaptive response to loading in mice is associated with sex-related deficiencies in osteoblasts but no change in osteocytes, Journal of Bone and Mineral Research 29(8) (2014) 1859–1871.

[40] P. Milovanovic, E.A. Zimmermann, M. Hahn, D. Djonic, K. Puschel, M. Djuric, M. Amling, B. Busse, Osteocytic Canalicular Networks: Morphological Implications for Altered Mechanosensitivity, ACS Nano 7(9) (2013) 7542–7551.

[41] K. Kobayashi, H. Nojiri, Y. Saita, D. Morikawa, Y. Ozawa, K. Watanabe, M. Koike, Y. Asou, T. Shirasawa, K. Yokote, Mitochondrial superoxide in osteocytes perturbs canalicular networks in the setting of age-related osteoporosis, Sci Rep 5(1) (2015) 1–11.

[42] E.S. Kurland, C.J. Rosen, F. Cosman, D. McMahon, F. Chan, E. Shane, R. Lindsay, D. Dempster, J.P. Bilezikian, Insulin-like growth factor-I in men with idiopathic osteoporosis, J. Clin. Endocrinol. Metab. 82(9) (1997) 2799–2805.

[43] Y. Pernow, E.M. Hauge, K. Linder, E. Dahl, M. Saaf, Bone Histomorphometry in Male Idiopathic Osteoporosis, Calcified Tissue International 84(6) (2009) 430–438.

[44] B.Y. Reed, J.E. Zerwekh, K. Sakhaee, N.A. Breslau, F. Gottschalk, C.Y.C. Pak, Serum Igf-1 Is Low And Correlated With Osteoblastic Surface In Idiopathic Osteoporosis, Journal of Bone and Mineral Research 10(8) (1995) 1218–1224.

[45] P.J. Marie, M.C. Devernejoul, D. Connes, M. Hott, Decreased Dna-Synthesis By Cultured Osteoblastic Cells In Eugonadal Osteoporotic Men With Defective Bone-Formation, J. Clin. Invest. 88(4) (1991) 1167–1172.

[46] S. Khosla, Idiopathic osteoporosis - Is the osteoblast to blame?, J. Clin. Endocrinol. Metab. 82(9) (1997) 2792–2794.

[47] J.A. Kanis, L.J. Melton III, C. Christiansen, C.C. Johnston, N. Khaltaev, The diagnosis of osteoporosis, Journal of bone and mineral research 9(8) (1994) 1137–1141.

[48] G. Karaguzel, M.F. Holick, Diagnosis and treatment of osteopenia, Reviews in endocrine and metabolic disorders 11(4) (2010) 237–251.

[49] G.M. Blake, I. Fogelman, The role of DXA bone density scans in the diagnosis and treatment of osteoporosis, Postgraduate medical journal 83(982) (2007) 509–517.

[50] A. El Maghraoui, C. Roux, DXA scanning in clinical practice, QJM: An International Journal of Medicine 101(8) (2008) 605–617.

[51] T. Davis, D. Healy, A. Bubeck, R. Walker, Stress concentrations around voids in three dimensions: The roots of failure, J. Struct. Geol. 102 (2017) 193–207.

[52] T.M. Skerry, L. Bitensky, J. Chayen, L.E. Lanyon, Early strain-related changes in enzyme activity in osteocytes following bone loading in vivo, J Bone Miner Res 4(5) (1989) 783–8.

[53] F.M. Lambers, G. Kuhn, F.A. Schulte, K. Koch, R. Muller, Longitudinal assessment of in vivo bone dynamics in a mouse tail model of postmenopausal osteoporosis, Calcif Tissue Int 90(2) (2012) 108–19.

[54] S.D. Badilatti, P. Christen, I. Parkinson, R. Müller, Load-adaptive bone remodeling simulations reveal osteoporotic microstructural and mechanical changes in whole human vertebrae, Journal of biomechanics 49(16) (2016) 3770–3779.

[55] M.R. Rubin, D.H. Schussheim, C.A.M. Kulak, E.S. Kurland, C.J. Rosen, J.P. Bilezikian, E. Shane, Idiopathic osteoporosis in premenopausal women, Osteoporosis International 16(5) (2005) 526–533.

[56] M.L. Gourlay, S.A. Brown, Clinical considerations in premenopausal osteoporosis, Arch. Intern. Med. 164(6) (2004) 603–614.

[57] K.E. Bainbridge, M. Sowers, X.H. Lin, S.D. Harlow, Risk factors for low bone mineral density and the 6-year rate of bone loss among premenopausal and perimenopausal women, Osteoporosis International 15(6) (2004) 439–446.

[58] C.A.M. Kulak, D.H. Schussheim, D.J. McMahon, E. Kurland, S.J. Silverberg, E.S. Siris, J.P. Bilezikian, E. Shane, Osteoporosis and low bone mass in premenopausal and perimenopausal women, Endocrine Practice 6(4) (2000) 296–304.

[59] M.R. Sowers, B. Shapiro, M.A. Gilbraith, M. Jannausch, Health And Hormonal Characteristics Of Premenopausal Women With Lower Bone Mass, Calcified Tissue International 47(3) (1990) 130–135.

[60] L. Karim, J. Moulton, M. Van Vliet, K. Velie, A. Robbins, F. Malekipour, A. Abdeen, D. Ayres, M.L. Bouxsein, Bone microarchitecture, biomechanical properties, and advanced glycation end-products in the proximal femur of adults with type 2 diabetes, Bone 114 (2018) 32–39.

[61] E.M. Lewiecki, P.D. Miller, Skeletal effects of primary hyperparathyroidism: bone mineral density and fracture risk, J. Clin. Densitom. 16(1) (2013) 28–32.

[62] K.J. Martin, K. Olgaard, J.W. Coburn, G.M. Coen, M. Fukagawa, C. Langman, H.H. Malluche, J.T. McCarthy, S.G. Massry, O. Mehls, Diagnosis, assessment, and treatment of bone turnover abnormalities in renal osteodystrophy, American journal of kidney diseases 43(3) (2004) 558–565.

[63] B. Hesse, M. Langer, P. Varga, A. Pacureanu, P. Dong, S. Schrof, N. Männicke, H. Suhonen, C. Olivier, P. Maurer, Alterations of mass density and 3D osteocyte lacunar properties in bisphosphonate-related osteonecrotic human jaw bone, a synchrotron μCT study, PloS one 9(2) (2014) e88481.

[64] J.R. Furst, L.C. Bandeira, W.W. Fan, S. Agarwal, K.K. Nishiyama, D.J. McMahon, E. Dworakowski, H. Jiang, S.J. Silverberg, M.R. Rubin, Advanced Glycation Endproducts and Bone Material Strength in Type 2 Diabetes, J Clin Endocrinol Metab 101(6) (2016) 2502–10.

[65] T. Rodic, E.M. Wölfel, P. Milovanovic, I.A. Fiedler, D. Cvetkovic, K. Jähn, M. Amling, J. Sopta, S. Nikolic, V. Zivkovic, Bone quality analysis of jaw bones in individuals with type 2 diabetes mellitus— post mortem anatomical and microstructural evaluation, Clinical Oral Investigations (2021) 1–24.

